# Subclinical Vascular Dysfunction in Mothers of Children with Anorectal Malformations: A Multiomics Study

**DOI:** 10.1101/2025.06.29.25330492

**Authors:** Aditi Kaloni, Rakesh S Joshi, Jaishri Ramji, Shrinal Vasa, Swayamprabha Samantaray, Khyati Sharma, Vaibhav Bhatt, Anupama Modi, Bhavin Parekh

## Abstract

**Background:** Anorectal malformations (ARMs) are rare birth defects with established maternal risk factors. However, the molecular mechanisms linking these risk factors to ARM development remain unclear. Maternal vascular dysfunction may serve as a unifying mechanistic pathway.

**Objective:** To investigate whether mothers of ARM-affected children exhibit molecular signatures of vascular dysfunction using integrated microbiome and metabolomic profiling.

**Methods:** Mothers of ARM children (cases; n = 10) and matched healthy controls (n = 10), recruited more than one year postpartum, were analyzed using 16S rRNA sequencing and untargeted LC-MS-based serum metabolomics. Microbial diversity was assessed using Shannon and Chao1 indices; compositional differences were evaluated by PERMANOVA across multiple distance metrics. Metabolites were analyzed using univariate and multivariate methods with FDR correction (q < 0.05).

**Results:** Cases exhibited elevated Firmicutes:Bacteroidetes ratios (70.1% vs 51.6%) and significant depletion of Olsenella (6.2% vs 16.4%, p < 0.05), a genus associated with cortisol metabolism. Metabolomic profiling revealed 174 significantly altered serum metabolites (p < 0.05, fold change >1.5). Among these, reduced D-glutamic acid, elevated thromboxane A , and decreased hexacosanoic acid emerged as key discriminants, mapping to pathways involved in oxidative stress, vasoconstriction, and peroxisomal dysfunction.

**Conclusions:** This pilot study reveals maternal microbiome-metabolome signatures suggestive of subclinical vascular dysfunction, resembling aspects of pre-eclampsia, in mothers of children with ARMs. These findings support the hypothesis that maternal vascular health influences ARM risk and warrant validation in larger, prospective cohorts.

**Graphical Abstract:** 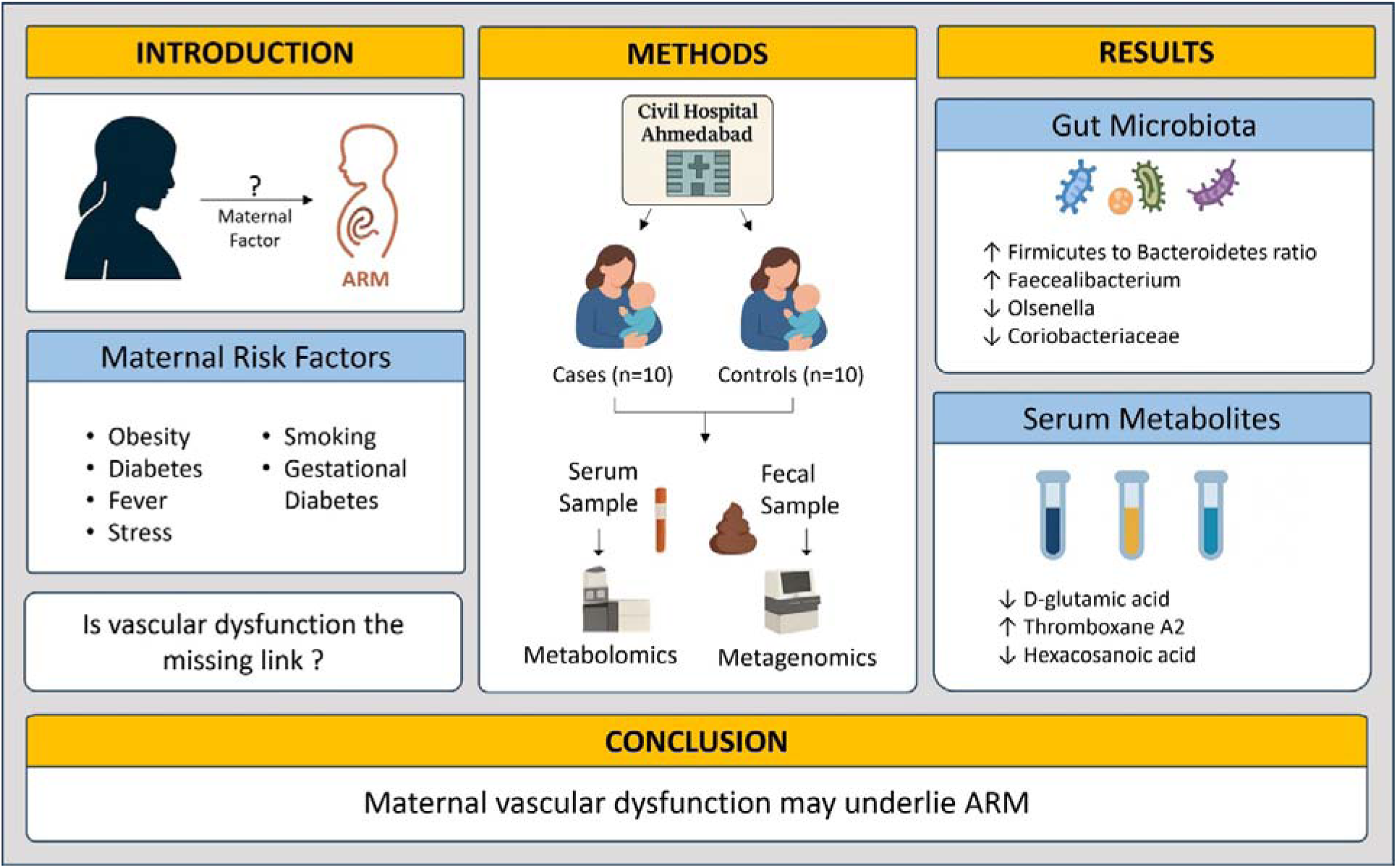

## 1. Introduction

Anorectal malformations (ARMs) are rare birth defects of the anus that obstruct stool evacuation(Levitt & Peña, 2007; Xie et al., 2025). ARMs affect around 3.11 cases per 10,000 live births (Xie et al., 2025). ARMs occur when the embryonic cloaca fails to separate into distinct urinary, genital, and digestive systems during the first trimester (De Blaauw et al., 2024; Gupta et al., 2014; Thomas, 2020; Zwink et al., 2011). Clinically, ARMs commonly present as either no anal opening (atresia) or unusual connections (fistulae) between the rectum and other nearby structures (e.g., skin)(De Blaauw et al., 2024; Levitt & Peña, 2007). Despite advanced anorectal reconstruction surgery, many children struggle with lifelong problems like poor bowel control, urogenic bladder, urinary tract infections, and kidney failure(De Blaauw et al., 2024; Gupta et al., 2014; Yang et al., 2020). Consequently, even with their relative rarity, ARMs impose substantial long-term physical, economic, and emotional burdens on affected individuals and families (De Blaauw et al., 2024).

Understanding the causes of ARMs remains challenging (Gupta et al., 2014). Some familial and syndromic cases, such as Townes-Brocks and Currarino syndromes, clearly involve genetic factors (De Blaauw et al., 2024; Khanna et al., 2018). Research also shows that ARM pathogenesis involves major genes impacting Shh, WNT, and Fgf signaling pathways and their downstream effectors (e.g., Gli2, BMP7, Fgf10, Wnt5a) (Draaken et al., 2012; Khanna et al., 2018; Yang et al., 2020). Yet nearly half of all cases occur sporadically, suggesting strong interactions between genetic susceptibility and environmental exposures (De Blaauw et al., 2024; Draaken et al., 2012; Dworschak et al., 2017; Khanna et al., 2018). Among environmental factors, maternal health is particularly important—with factors like obesity, diabetes, gestational diabetes, smoking, and fevers early in pregnancy increasing ARM risk(De Blaauw et al., 2024; Van Rooij et al., 2010; Wijers et al., 2013). Exactly how these maternal factors lead to ARMs remains unclear.

Closer examination of maternal risk factors reveals a cogent hypothesis: vascular dysfunction. Maternal obesity and diabetes, including gestational diabetes, are linked to vascular endothelial dysfunction (McElwain et al., 2020; Moraes et al., 2021). Febrile illnesses cause vascular damage and placental infarction (Mamtha K. et al., 2023). Cigarette smoking damages the vascular system through endothelial dysfunction, oxidative stress, inflammation, and thrombosis(Hahad et al., 2023). The temporal overlap between anorectal differentiation and maternal vascular adaptation during the first trimester further strengthens this proposal. Between gestational weeks 4 and 8, the embryonic cloaca must divide into separate genitourinary and intestinal systems—precisely when maternal vascular remodeling and placental development are most active (Boeldt & Bird, 2016; Gupta et al., 2014; Reijnders et al., 2021; Thomas, 2020; Zwink et al., 2011). Since uteroplacental blood circulation is essential for embryonic and fetal development, this overlap suggests that disruptions in maternal vascular health impact embryonic anorectal development(Radford et al., 2023; Reijnders et al., 2021; Weckman et al., 2019). Preeclampsia, especially early-onset type (<34 weeks of gestation), reinforces this vascular connection(Boeldt & Bird, 2016; Shaw et al., 2024). Marked by defective placental invasion and impaired maternal artery remodeling, it causes placental insufficiency and increases the risk of birth defects—including defects of the heart and anorectum(Ferreira et al., 2022; Radford et al., 2023; Shaw et al., 2024; Weber et al., 2018; Wijers et al., 2013). However, direct evidence linking maternal vascular dysfunction to ARM pathogenesis remains unexplored.

To uncover molecular evidence of this vascular link, we conducted the first multiomics pilot study comparing mothers of ARM-affected children with controls. We profiled gut microbiota and serum metabolomes as these systems reflect systemic vascular health and can reveal subclinical dysfunction. This approach relies on the fact that maternal metabolic and microbial profiles remain relatively stable postpartum, providing insights into periconceptional physiology.

## 2. Methods

### 2.1 Study Design and Settings

We conducted a single-center, retrospective, cross-sectional case-control pilot study from December 2023 to May 2024 at the Department of Pediatric Surgery, Civil Hospital, Ahmedabad, Gujarat, India. The study aimed to compare the gut microbiota composition and serum metabolomic profiles of mothers of children with anorectal malformations (ARMs) (cases; n□=□10) with those of mothers of healthy children (controls; n□=10).

Participants were recruited using purposive random sampling to ensure balanced representation across groups (Figure 1). Given the rarity of ARM, conducting a prospective cohort study during pregnancy would require an impractically large sample size. Therefore, we assessed mothers after one year postpartum, a time point at which maternal gut microbiota and dietary patterns have been shown to stabilize and return to pre-pregnancy states, allowing for a valid approximation of baseline maternal health status (T. Wang et al., 2021).

**Figure 1:**
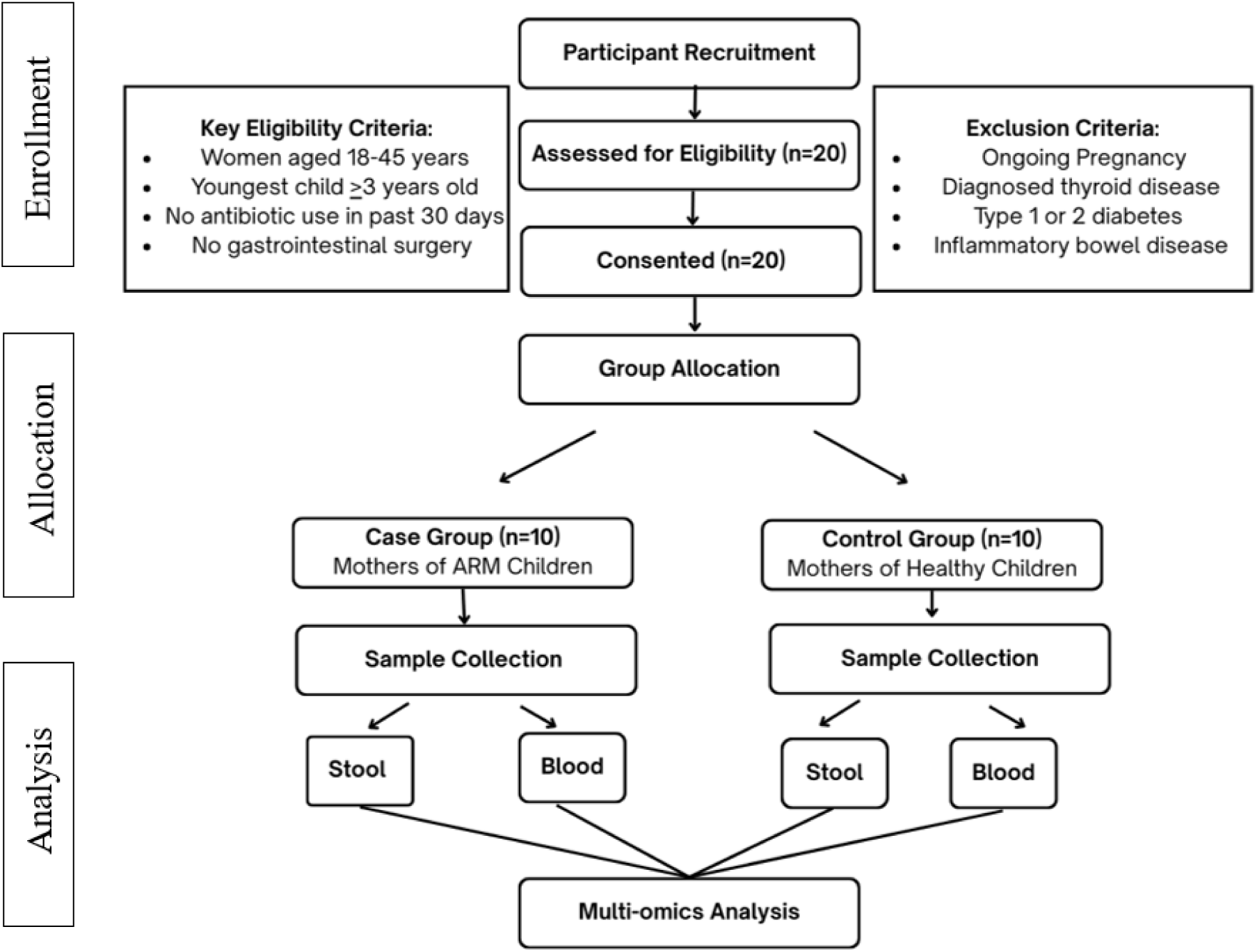
Flowchart illustrating the enrollment, allocation, and analysis process of the study participants

#### Sample Size

This pilot study was powered to detect large effect sizes (Cohen’s d ≥1.2) with 80% power at α=0.05 (Cohen, 2009; Lakens, 2013). The sample size is in line with exploratory multiomics studies, where pilot studies of 10-20 samples per group are considered sufficient for hypothesis generation and effect size estimation for future confirmatory studies (Billoir et al., 2015; Nyamundanda et al., 2013; Tarazona et al., 2020).

### 2.2 Participants and Recruitment

#### Inclusion and Exclusion Criteria

Cases comprised mothers whose children had received a clinical diagnosis of any anorectal malformation confirmed by pediatric surgeons. Controls consisted of mothers whose children exhibited no congenital anomalies or chronic health conditions. All participants were females aged between 18 and 45 years, capable of providing informed consent, and whose youngest child was at least three years old, ensuring sufficient post-birth physiological recovery. Exclusion criteria applied uniformly to both groups included: current pregnancy, diagnosed thyroid disorders, type 1 or type 2 diabetes mellitus, antibiotic use within the past 30 days, a history of gastrointestinal surgery, inflammatory bowel diseases, and chronic usage of medications known to significantly affect gut microbiota.

#### Recruitment Strategy

Given the rarity of ARMs and the limited availability of eligible participants during the study period, formal matching between cases and controls (e.g., by age or BMI) was not feasible. Cases were recruited through the pediatric surgery outpatient clinic at Civil Hospital, Ahmedabad, primarily from families attending follow-up visits. Controls were enrolled from the same geographic region. This strategy, a potential source of selection bias, was considered a necessary and pragmatic approach given the rarity of the condition and the resource constraints inherent to a pilot study.

### 2.3 Ethical Considerations

The study received approval from the Institutional Ethics Committee of B.J. Medical College & Civil Hospital (EC/Approval/104/2023/04/12/2023). All participants provided written informed consent. The consent process included verbal explanation in their preferred language—Gujarati, Hindi, or English. Participants retained withdrawal rights throughout the study. Complete confidentiality was maintained for all data.

### 2.4 Sample Collection and Processing

Fecal samples were collected in sterile containers using standardized collection kit and stored at-80°C within 2 hours of collection. Venous blood (10 mL) was drawn into serum separator tubes, allowed to clot for 60 minutes at room temperature, then centrifuged at 1300×g for 5 minutes. Serum aliquots (∼0.5 mL) were then stored at -80°C. Sample processing occurred at Gujarat Biotechnology Research Centre (GBRC) and Gujarat Technological University (GTU).

### 2.5 Metagenomic DNA Extraction and Sequencing

Metagenomic DNA was extracted from 250 mg fecal samples using the HiPurA Stool DNA Purification Kit (HiMedia) following the manufacturer’s protocol. DNA purity and concentration were evaluated using NanoDrop spectrophotometry (Thermo Fisher Scientific). Only samples meeting quality criteria of A260/280 ratios >1.8 and A260/230 ratios between 2.0-2.2 were included in downstream analyses.

The V3–V4 hypervariable regions of the bacterial 16S rRNA gene were amplified using universal primers 341F (5′-CCTACGGGNGGCWGCAG-3′) and 785R (5′-GACTACHVGGGTATCTAATCC-3′) appended with Illumina overhang adapters. PCR was performed using KAPA HiFi HotStart ReadyMix. Cycling conditions included initial denaturation at 95°C for 3 minutes, followed by 25 cycles of 95°C for 30 seconds, 55°C for 30 seconds, and 72°C for 30 seconds. Final extension occurred at 72°C for 5 minutes.

Library preparation followed standard protocols. Amplicons were purified using AMPure XP beads and quantified using Qubit fluorometry. After normalization, samples were indexed using the Nextera XT Index Kit. Sequencing was performed on the Illumina MiSeq platform using 2×300 bp paired-end chemistry with 5% PhiX spike-in.

### 2.6 Metagenomic Data Analysis

Raw sequences underwent quality control using QIIME2 (version 2024.2). Sequences were trimmed to a fixed length of 250 bp, and low-quality reads were filtered. Chimeric sequences were identified and removed using VSEARCH. Amplicon sequence variants (ASVs) were generated using the Deblur plugin. Taxonomic classification was performed using the GreenGenes database (v13_8) at 99% similarity. ASVs with fewer than 10 total reads were removed from downstream analyses.

Diversity analyses followed standard practices. Alpha diversity was calculated using Shannon and Chao1 indices. Beta diversity employed Bray-Curtis, Jaccard, weighted and unweighted UniFrac distance matrices. Rarefaction depth was set to 1000 reads per sample based on rarefaction curve analysis, where diversity metrics plateaued. Statistical significance of beta diversity differences was tested using PERMANOVA with 999 permutations.

### 2.7 Untargeted Serum Metabolomics

Sample preparation followed protein precipitation protocols. Serum samples (100 μL) were treated with 900 μL ice-cold acetonitrile, vortexed for 30 seconds, and centrifuged at 12,000×g for 15 minutes at 4°C. The resulting supernatants were transferred to autosampler vials.

We employed high-resolution mass spectrometry for metabolite detection. Metabolites were analyzed using the Agilent 6545XT Advanced Bio LC-MS Q-TOF system in positive electrospray ionization mode (ESI+). The mass range covered 50-1200 m/z. Chromatographic separation utilized a C18 column (2.1×100 mm, 1.7 μm) maintained at 35°C with a flow rate of 0.200 mL/min.

Mobile phase composition was optimized for metabolite coverage. Phase A consisted of 0.1% formic acid in water. Phase B contained 0.1% formic acid in acetonitrile. Total run time was of 25 min with gradient varying from 1% to 95%. Quality control measures ensured data reliability.

Pooled quality control samples were analyzed at the end of the batch to monitor instrument performance. Blank samples assessed background contamination. Only metabolites with coefficient of variation <30% in QC samples were retained for analysis. This stringent filtering maintained data quality while preserving biological signal. Raw data underwent systematic processing for peak detection and metabolite identification. Files were converted to mzML format using ProteoWizard software. Peak picking was performed using the centWave algorithm, followed by retention time correction and grouping of features across samples. The CAMERA package was used for peak annotation, including isotopes and adducts. Metabolite identification was carried out by matching accurate mass (±5 ppm tolerance) against the HMDB database.

Data were imported into MetaboAnalyst v6.0 for statistical analysis. Median normalization corrected for systematic variations between samples. Auto-scaling was applied prior to multivariate analysis. Statistical analysis combined univariate and multivariate approaches. Multivariate analysis included PCA for data visualization and OPLS-DA for group discrimination. Univariate analysis employed Welch’s t-test for between-group comparisons. Significant metabolites were defined by three criteria: p<0.05, |fold change|>1.5, and VIP>1.0. Multiple testing correction addressed the problem of inflated Type I error rates. The Benjamini-Hochberg procedure controlled the false discovery rate (FDR) at q<0.05. Heatmaps were used for visualization of differential metabolite patterns, and pathway enrichment analyses was conducted using KEGG and disease-related pathway databases to interpret biological relevance.

### 2.8 Statistical Analysis

Continuous variables were tested for normality using the Shapiro–Wilk test. Non-normally distributed data were compared using the Mann–Whitney U test and reported as median with interquartile range (IQR). Categorical variables were analyzed using Fisher’s exact test. Beta diversity differences were assessed using PERMANOVA with 999 permutations across multiple distance metrics. Metabolomics analysis combined Welch’s t-test for univariate comparisons and multivariate methods, including principal component analysis (PCA) and orthogonal partial least squares-discriminant analysis (OPLS-DA). Models were validated using cross-validation and permutation testing. FDR correction (q < 0.05) was applied to all univariate analyses to control for multiple testing. Microbiome–metabolome integration used Spearman’s rank correlation. Correlation matrices were FDR-adjusted, and only associations with q < 0.05 were deemed significant.

## 3. Results

### 3.1 Study Population

Twenty mother-child dyads were enrolled and equally divided between cases and controls. Cases were younger (median 25 ± 4.75 vs. 32 ± 2 years, Mann-Whitney U test, p=0.0047) (Table 1). Age distributions of children were similar (2.1 ± 1.225 vs. 2.25 ± 1 years, Mann-Whitney U test, p=0.642) (Table 1). Sex ratios favored males in both groups (70% ARM vs. 60% controls, Fisher’s exact test, p=1.000). All births were full-term deliveries. Delivery and health patterns differed between groups. ARM cases had higher rates of vaginal delivery (70% vs. 20%, Fisher’s exact test, p=0.16), though this did not reach statistical significance. Comorbidities occurred exclusively in ARM-affected children (40% vs. 0%, Fisher’s exact test, p=0.414). These differences, while notable, must be interpreted cautiously given small sample sizes.

**Table 1:** Characteristics of Children and Delivery Details.

| Characteristics | Case Frequency | Case Percentage | Control Frequency | Control Percentage | p-value |
| --- | --- | --- | --- | --- | --- |
| Child Age (Median ± IQR) | 2.1 ± 1.225 | N/A | 2.25 ± 1 | N/A | 0.6421486 |
| Child gender - Female | 3 | 30% | 4 | 40% | 1 |
| Child gender - Male | 7 | 70% | 6 | 60% | 1 |
| Delivery type - Normal | 7 | 70% | 2 | 20% | 0.16 |
| Delivery type - C-section | 3 | 30% | 8 | 80% | 0.2 |
| Delivery maturity - Pre term | 0 | 0% | 0 | 0% | 1 |
| Delivery maturity - Full term | 10 | 100% | 10 | 100% | 1 |
| Comorbidity of ARM child - Yes | 4 | 40% | 0 | 0% | 0.117 |
| Comorbidity of ARM child - No | 6 | 60% | 10 | 100% | 0.414 |

Values for numerical variables are presented as median ± IQR. The Chi-square exact test was used for qualitative and categorical data, and values are expressed as number (percentage).

### 3.2 Maternal Characteristics and Pregnancy History

Maternal characteristics revealed several group differences. Cases reported higher rates of pregnancy-related stress (70% vs. 10%, Fisher’s exact test, p=0.039) (Table 3). Pregnancy complications occurred more frequently in cases (40% vs. 10%, Fisher’s exact test, p=0.335). Miscarriages were reported exclusively among cases (20%, Fisher’s exact test, p=0.485).Dietary patterns showed uniform vegetarianism among cases compared to 70% among controls (Fisher’s exact test, p=0.523) (Table 2). Paternal substance use rates were higher in the ARM group (70% vs. 20%, Fisher’s exact test, p=0.127). However, most maternal characteristic differences did not achieve statistical significance, likely due to limited sample size.

**Table 2:** Paternal History and Dietary Factors.

| Characteristics | Case Frequency | Case Percentage | Control Frequency | Control Percentage | p-value |
| --- | --- | --- | --- | --- | --- |
| Addiction of father<br>- Yes | 7 | 70% | 2 | 20% | 0.1273861 |
| Addiction of father<br>- No | 3 | 30% | 8 | 80% | 0.1551874 |
| Diet - Vegetarian | 10 | 100% | 7 | 70% | 0.5231071 |
| Diet - Non-vegetarian | 0 | 0% | 3 | 30% | 0.2307692 |

Chi-square exact test was used for qualitative and categorical data, and values are expressed as number (percentage).<sup>\*</sup> Asterisks indicate statistical significance ( $p < 0.05$ ).

**Table 3:**
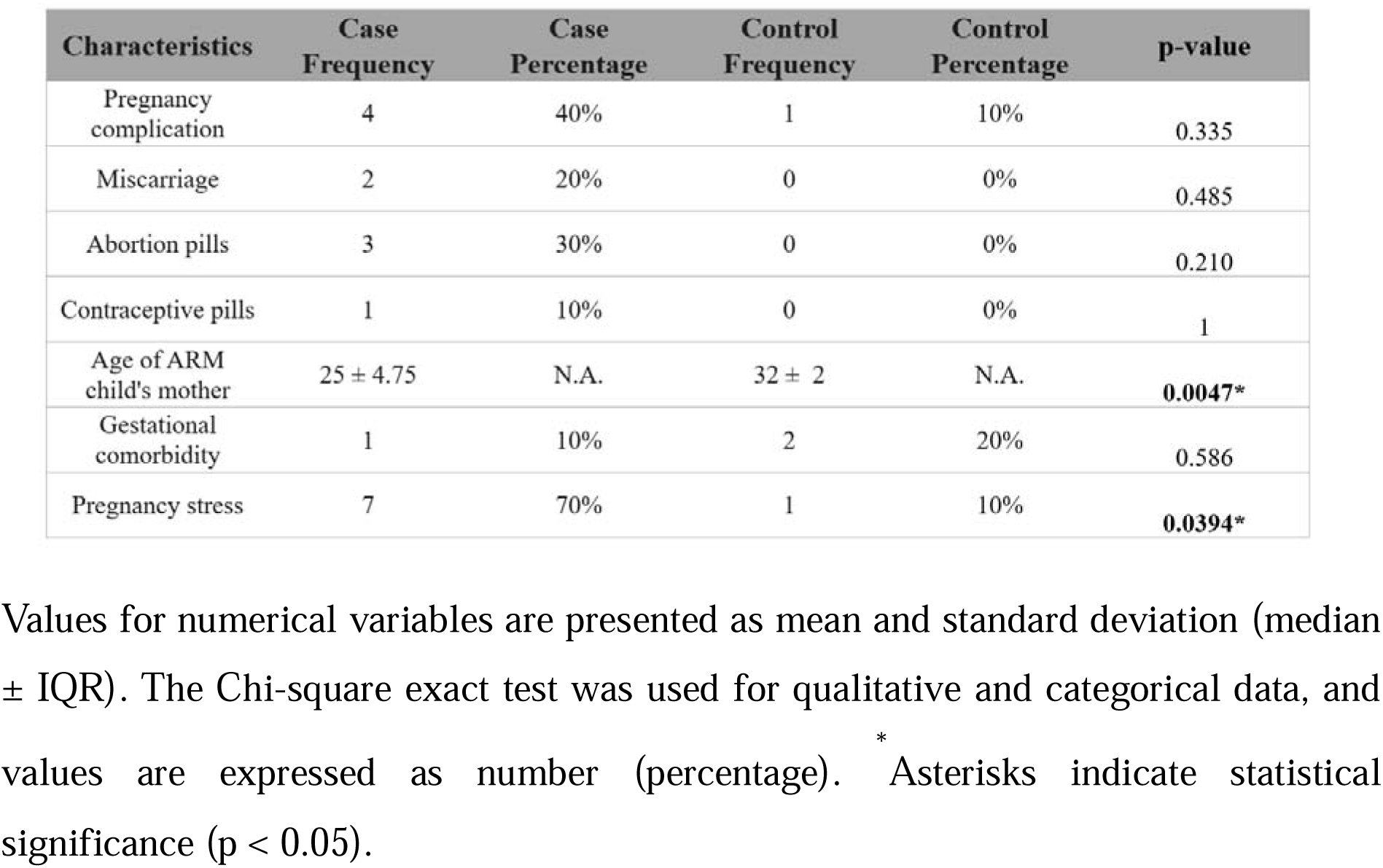
Pregnancy related characteristics.

### 3.3 Metagenomic Analysis Sequencing Quality and Coverage

Sequencing generated 9,852,532 paired-end reads with a median of 359,802 reads per sample (range: 151,446–1,022,477) (Supplementary Table 1). Rarefaction curve analysis confirmed sufficient sequencing depth, with α-diversity metrics reaching asymptotic plateaus, indicating comprehensive microbial community coverage (Figure 2a).

**Figure 2:**
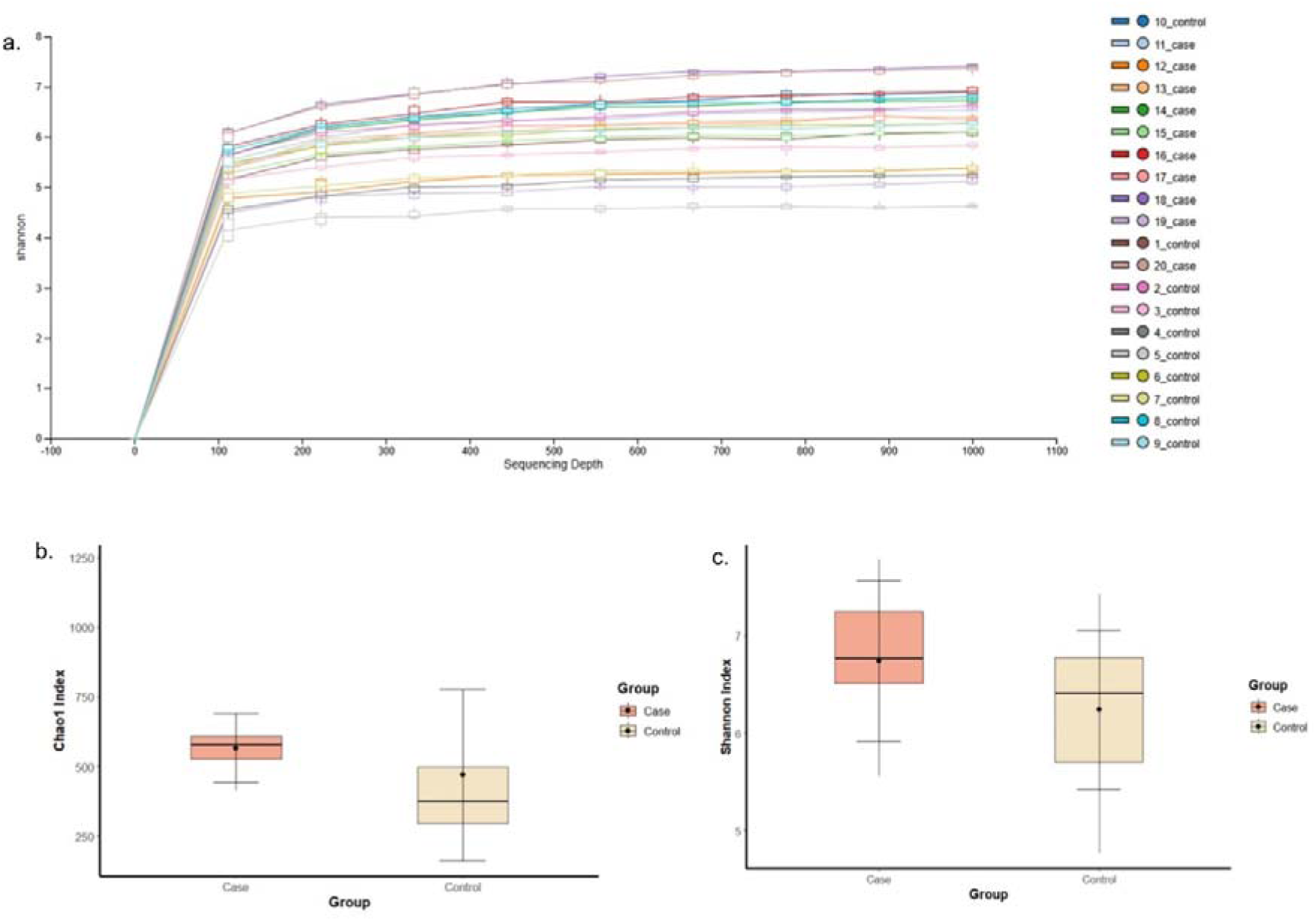
Microbiome communities from 20 mothers were sequenced using 16S rRNA. (a) Rarefaction curve showing alpha diversity (Shannon index) versus sequencing depth for case and control samples, with plateauing indicating sufficient sampling; (b) boxplot of the Chao1 index; and (c) boxplot of the Shannon index, both representing alpha diversity. Boxes show the interquartile range (IQR), with horizontal lines for medians and whiskers extending to the minimum and maximum values. No statistically significant differences were observed between groups (Chao1: *p* > 0.05, unpaired Student’s *t*-test; Shannon: *p* > 0.05, Mann-Whitney U test).

#### Microbial Diversity Assessment

α-diversity analyses revealed that cases had more complex gut bacterial communities than controls. Cases showed higher bacterial richness (median 580 vs. 350 species, pChao1>0.05) and more similar species distribution (Shannon diversity 6.8 vs. 6.4, pShannon>0.05). These consistent differences indicate that cases harbor distinct microbial ecosystems, though statistical significance was not reached in this pilot study. β-diversity analysis revealed compositional differences between groups. We found that microbial community structure differed significantly between cases and controls across multiple distance metrics: Bray-Curtis dissimilarity (PERMANOVA, R²=0.14, p<0.01) (Fig. 3a), Jaccard distance (PERMANOVA, R²=0.11, p<0.05) (Fig. 3b), unweighted UniFrac (PERMANOVA, R²=0.12, p<0.05) (Fig. 3c), and weighted UniFrac (PERMANOVA, R²=0.10, p<0.05) (Fig. 3d). These consistent patterns across phylogenetic and non-phylogenetic metrics suggest community structure differences.

**Figure 3:**
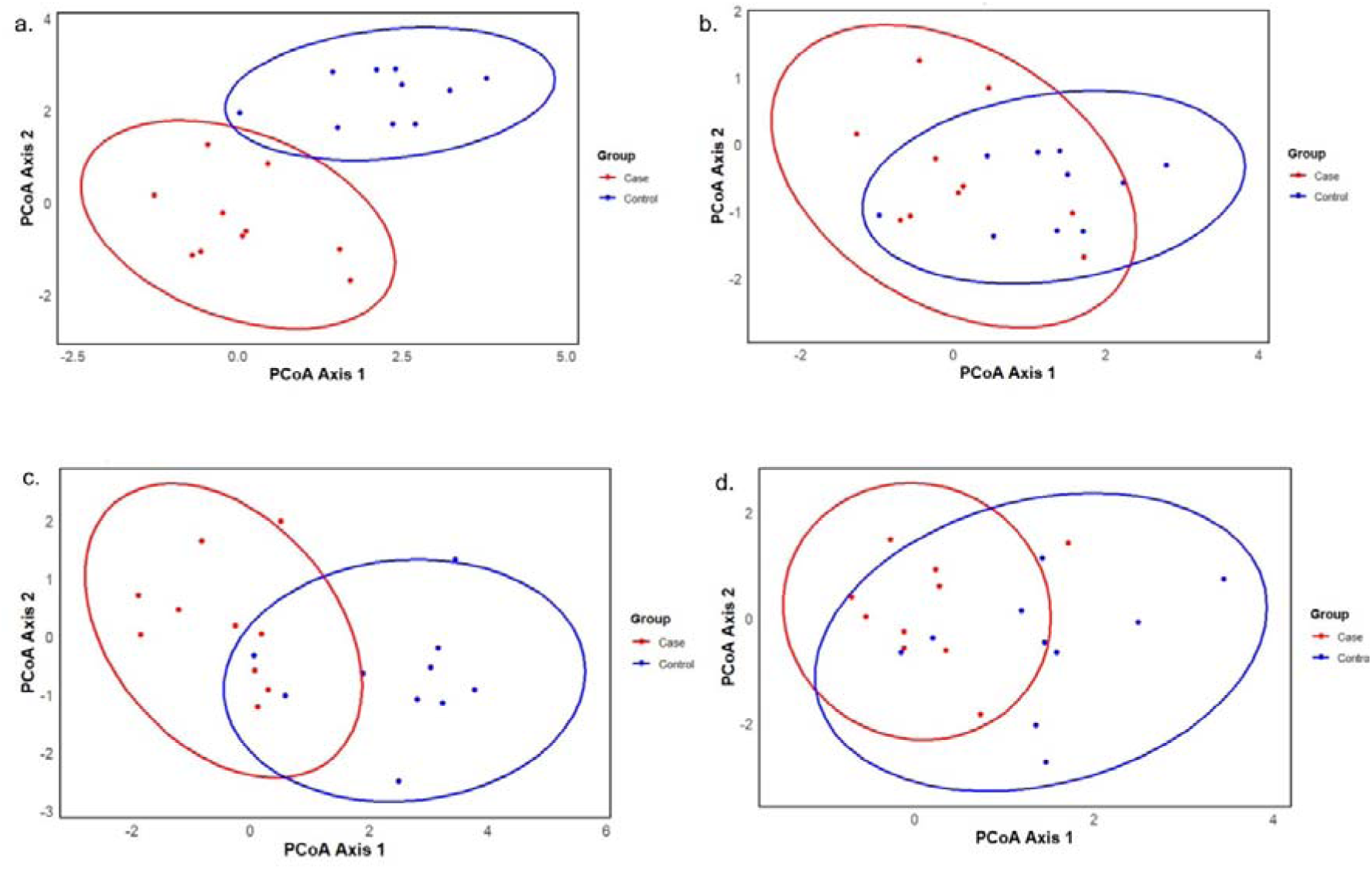
Principal coordinate analysis (PCoA) plots illustrating the beta-diversity of microbial communities across different samples. (a) Bray–Curtis; (b) Jaccard metrics; (c) unweighted UniFrac; and (d) weighted UniFrac plots illustrating group overlap with PERMANOVA results.

#### Taxonomic Profiling

Phylum-level analysis revealed shifts in major bacterial groups (Fig. 4a). Firmicutes dominated both groups but showed enrichment in cases (70.1% vs. 51.6%, Mann-Whitney U test, p<0.05). Conversely, Actinobacteria was depleted in cases (12.3% vs. 30.6%, Mann-Whitney U test, p<0.05). Bacteroidetes and Proteobacteria remained comparable between groups (Mann-Whitney U test, p>0.05 for both).

**Figure 4:**
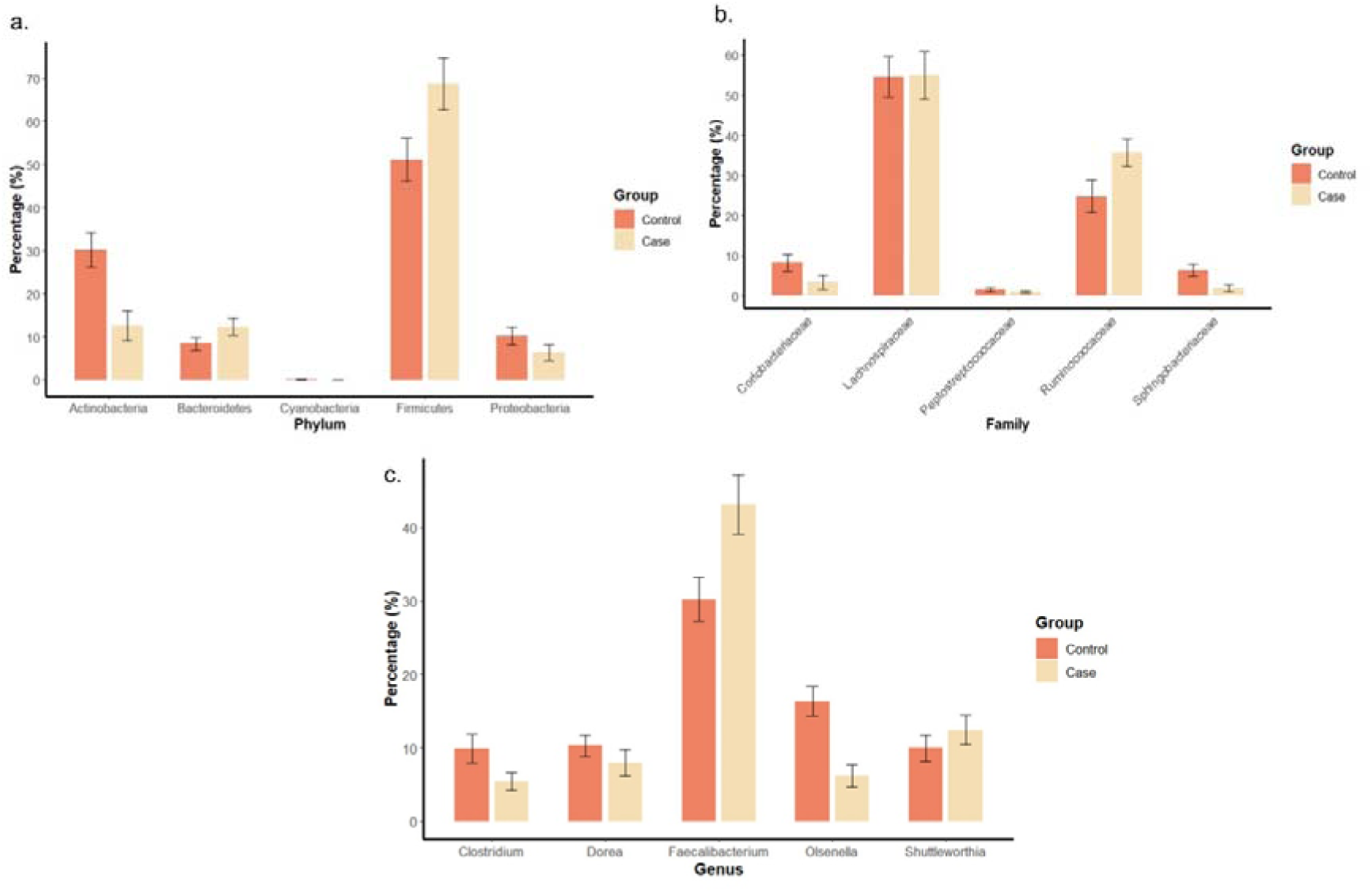
Taxonomic composition of gut microbiota at different taxonomic levels. (a) Relative abundance of bacterial phyla in case and control groups; (b) relative abundance of bacterial families in case and control groups; and (c) relative abundance of bacterial genera in case and control groups. Error bars show standard error. Statistical comparisons were performed using the Mann-Whitney U test. Significance notation: ns = not significant; * = *p* < 0.05.

Family-level differences were more pronounced (Fig. 4b). Cases exhibited higher Ruminococcaceae abundance (36.2% vs. 24.5%, Mann-Whitney U test, p<0.05). Lachnospiraceae remained stable at approximately 50% in both groups (Mann-Whitney U test, p>0.05). Controls showed greater prevalence of Corynebacteriaceae (10% vs. 5%, Mann-Whitney U test, p<0.05).

Genus-level analysis identified key discriminating taxa (Fig. 4c). Faecalibacterium was significantly enriched in cases (43.1% vs. 29.8%, Mann-Whitney U test, p<0.05). Olsenella showed marked depletion in cases (6.2% vs. 16.4%, Mann-Whitney U test, p<0.05). Clostridium was also reduced in cases (5.3% vs. 10.1%, Mann-Whitney U test, p<0.05). These changes represent substantial effect sizes despite small sample numbers.

### 3.4 Metabolomic Analysis

#### Differentially Expressed Metabolites in Serum Samples

Untargeted LC-MS-based metabolomics profiling of preprocessed serum samples yielded 1,054 distinct peaks (Supplementary Figure 1). Following rigorous quality control filtering, only metabolites that satisfied stringent inclusion criteria were retained for downstream analysis. A heatmap of the top 25 statistically significant differentially expressed metabolites revealed distinct group-wise clustering, (Fig. 5a).

**Figure 5:**
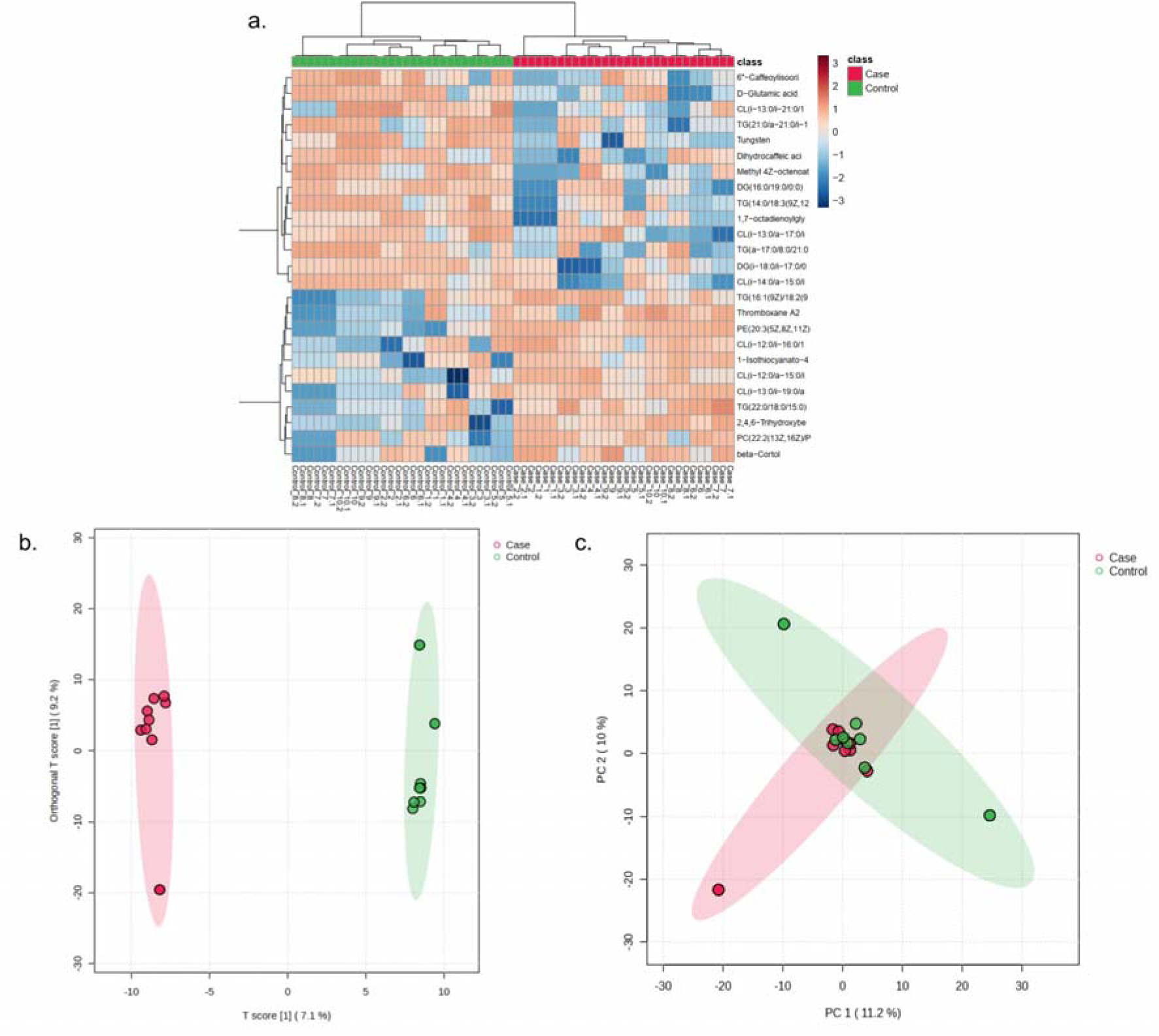
Metabolomic signatures distinguishing case and control groups. (a) Heatmap displaying the top 25 differentially expressed metabolites, based on statistical significance; (b) OPLS-DA score plot illustrating clear separation between case (red) and control (green) groups. The first two components explain 9.2% (PC1) and 7.1% (PC2) of the total variation; (c) PCA score plot showing the distribution of case (red) and control (green) samples, with PC1 accounting for 11.2% and PC2 for 10% of the

Multivariate analysis revealed distinct metabolomic signatures between cases and controls. Both Orthogonal Partial Least Squares Discriminant Analysis (OPLS-DA, R²=0.82, Q²=0.61) (Fig. 5b) and Principal Component Analysis (PCA, PC1=11.2%, PC2=10.0%) (Fig. 5c) showed clear group separation. These results indicate metabolomic differences despite small sample sizes.

We identified 369 metabolites with Variable Importance in Projection (VIP) scores >1.0, with the top 25 shown in (Fig. 6a). Comprehensive statistical analysis revealed 174 metabolites meeting our significance criteria (Welch’s t-test, p<0.05; fold change >1.5; VIP >1.0) (Fig. 6b) (Supplementary File 1). Among these differentially expressed metabolites (DEMs), 106 were upregulated and 100 were downregulated in cases compared to controls. A total of 174 DEMs were identified and selected for downstream analysis based on the overlap between univariate and multivariate analysis criteria and were visualized using a Venn diagram (Fig. 6c).

**Figure 6:**
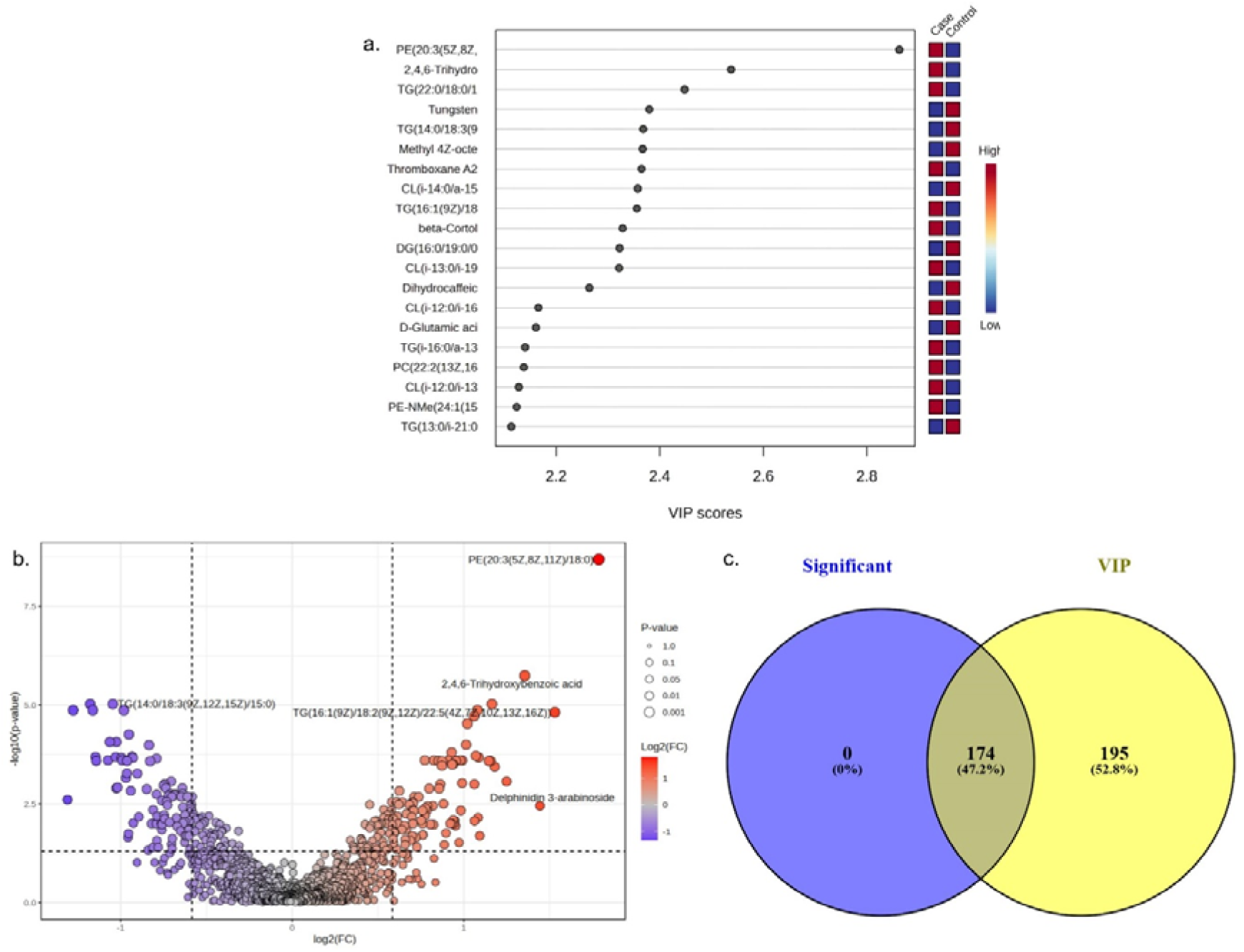
Integrated analysis of differential metabolites using multivariate and univariate statistical approaches. (a) VIP score plot showing the top 25 metabolites with VIP > 1.0, identified by multivariate analysis; (b) Volcano plot illustrating log2 fold change vs. –log10(p-value), highlighting metabolites with fold change > 1.5 and p-value < 0.05; (c) Venn diagram showing overlap of significant features identified by univariate (p < 0.05) and multivariate (VIP > 1.0) analyses.

#### Pathway Enrichment Analysis

Pathway enrichment analysis of the 174 DEMs revealed four significantly enriched metabolic pathways (Fig. 7a). These included D-amino acid metabolism (p<0.01), glycerophospholipid metabolism (p<0.05), arachidonic acid metabolism (p<0.05), and linoleic acid metabolism (p<0.05). These pathways suggest disrupted amino acid processing, membrane lipid metabolism, and inflammatory signaling in ARM mothers.

**Figure 7:**
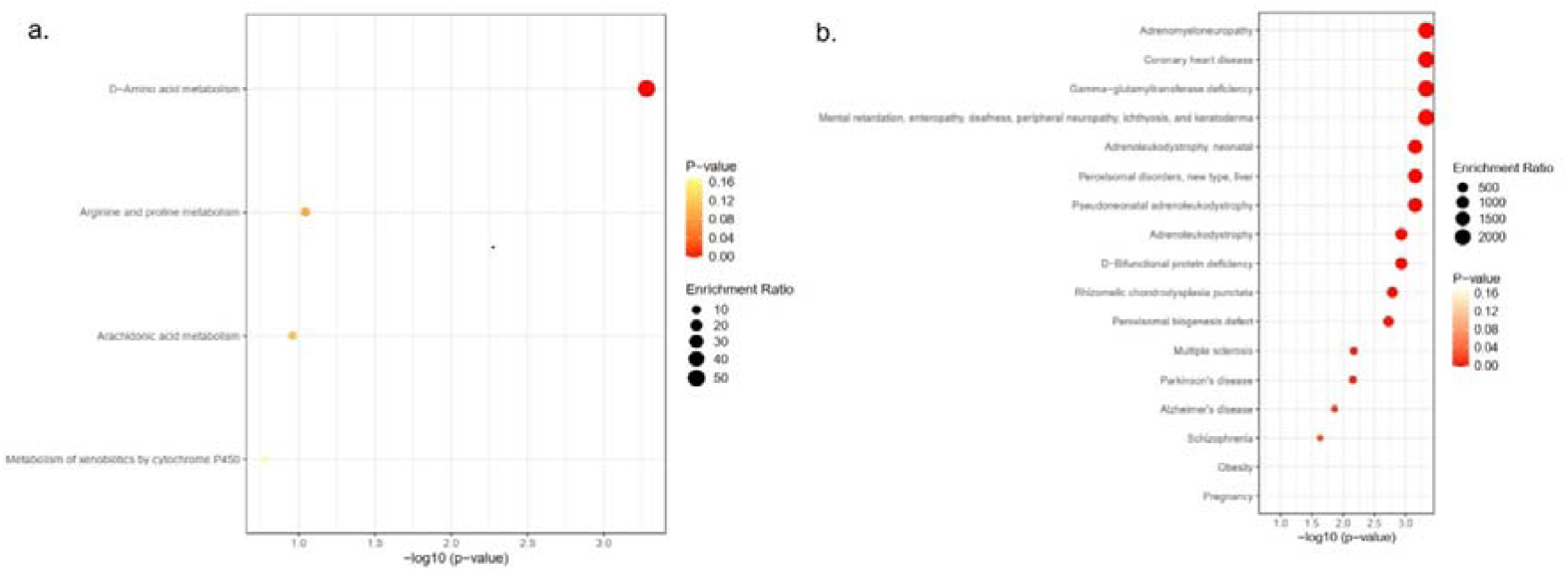
Enrichment analyses of differentially expressed metabolites (DEMs). (a) Pathway enrichment analysis of the 174 DEMs identified four significantly enriched metabolic pathways; (b) Disease enrichment analysis of the 174 DEMs identified significant associations with 15 diseases.

#### Disease-Based Enrichment Analysis

Disease enrichment analysis revealed significant associations between the 174 DEMs and 15 disease signatures (Fig. 7b). Top enriched conditions included coronary heart disease (FDR<0.05), adrenomyeloneuropathy (FDR<0.05), gamma-glutamyltransferase deficiency (FDR<0.05), adrenoleukodystrophy (FDR<0.05), and neonatal pseudoneonatal adrenoleukodystrophy (FDR<0.05). These associations suggest metabolic patterns consistent with vascular and oxidative stress disorders.

Four metabolites demonstrated statistically significant alterations between cases and controls (Table 4). D-glutamic acid exhibited significant reduction in cases compared to controls (Welch’s t-test, p<0.01) (Fig. 8a). Conversely, thromboxane A2 showed significant elevation in cases (Welch’s t-test, p<0.01) (Fig. 8b). Hexacosanoic acid was significantly decreased in cases (Welch’s t-test, p<0.05) (Fig. 8c), while tungsten levels were significantly reduced in the case group (Welch’s t-test, p<0.01) (Fig. 8d). These differentially abundant metabolites participated in multiple identified disease pathways, underscoring their potential significance in ARM-associated metabolic dysregulation (Table 4).

**Figure 8:**
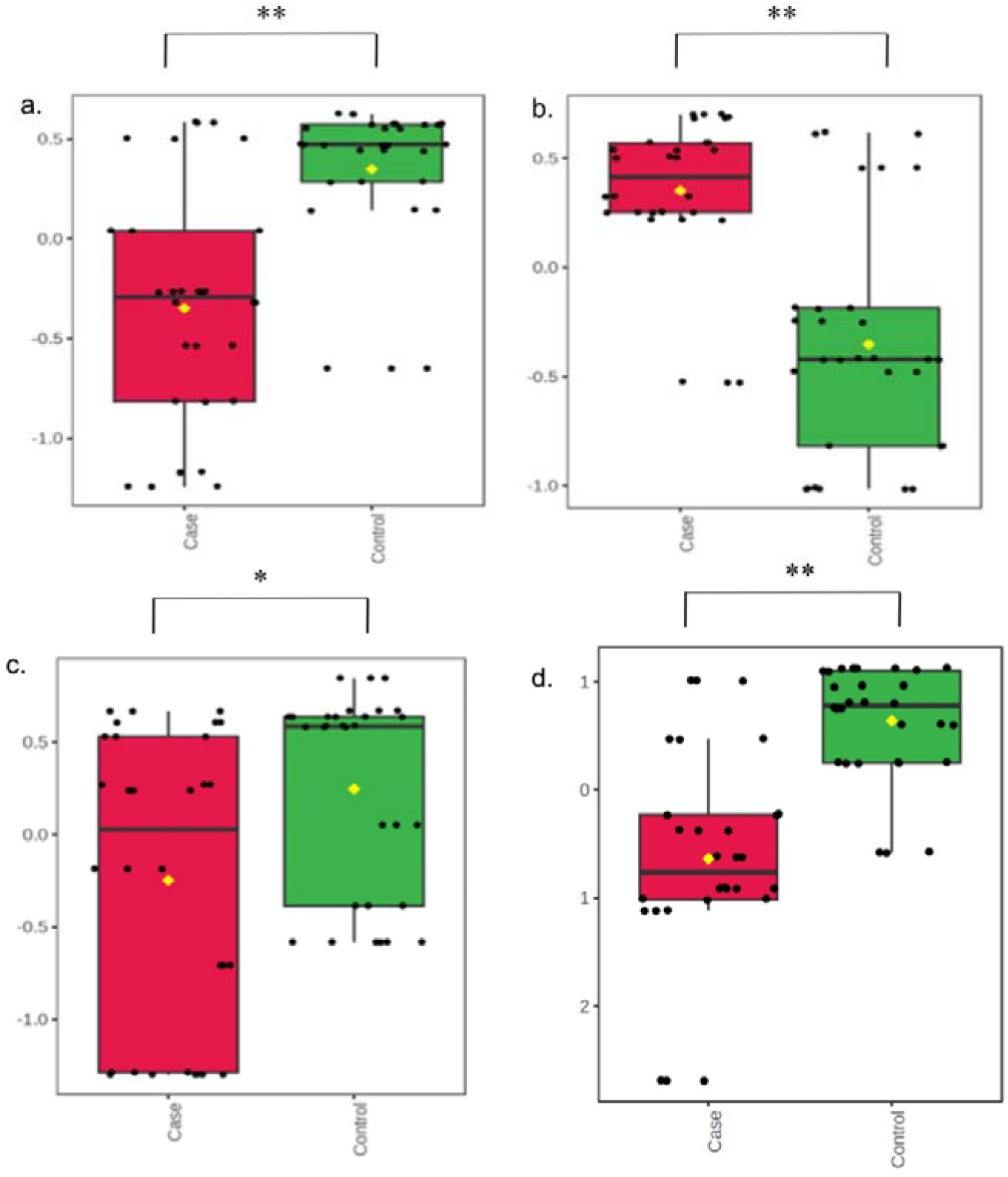
Top four significant pathways based on differentially expressed metabolites, with bar lengths indicating enrichment ratios. (a) Boxplot of D-glutamic acid; (b) thromboxane A2; (c) hexacosanoic acid; (d) and Tungsten, comparing case (red) and control (green) groups. *p*-value notation: ns = not significant; * = *p* < 0.05; ** = *p* < 0.01.

**Table 4:** Association of Various Diseases with Specific Metabolites and Statistical Significance.

| Sr. No. | Disease | p-value | FDR | Metabolite involved |
| --- | --- | --- | --- | --- |
| 1. | Adrenomyeloneuropathy | 4.67E-04 | 0.028 | Hexacosanoic acid |
| 2. | Coronary heart disease | 4.67E-04 | 0.028 | Thromboxane A2 |
| 3. | Gamma-glutamyltransferase deficiency | 4.67E-04 | 0.028 | D-Glutamic acid |
| 4. | Mental retardation, enteropathy, deafness, etc | 4.67E-04 | 0.028 | Hexacosanoic acid |
| 5. | Adrenoleukodystrophy, neonatal | 7.04E-04 | 0.028 | Hexacosanoic acid |
| 6. | Peroxisomal disorders, new type, liver | 7.04E-04 | 0.028 | Hexacosanoic acid |
| 7. | Pseudoneonatal adrenoleukodystrophy | 7.04E-04 | 0.028 | Hexacosanoic acid |
| 8. | Adrenoleukodystrophy | 0.00117 | 0.0363 | Hexacosanoic acid |
| 9. | D-Bifunctional protein deficiency | 0.00117 | 0.0363 | Hexacosanoic acid |
| 10. | Rhizomelic chondrodysplasia punctata | 0.00163 | 0.0457 | Hexacosanoic acid |
| 11. | Peroxisomal biogenesis defect | 0.00187 | 0.04575 | Hexacosanoic acid |
| 12. | Multiple sclerosis | 0.00675 | 0.15 | Tungsten |
| 13. | Parkinson's disease | 0.00698 | 0.15 | Tungsten |
| 14. | Alzheimer's disease | 0.0137 | 0.3 | Tungsten |
| 15. | Schizophrenia | 0.0234 | 0.436 | D-Glutamic acid |

**Table 5:** Table representing key metabolites identified through multivariate and univariate analysis.

| Metabolites | Retention time/sec | m/z | VIP | P value | FC value |
| --- | --- | --- | --- | --- | --- |
| Tungsten | 241.37 | 112.9842 | 2.380081794 | 2.92E-08 | 0.44205 |
| Thromboxane A2 | 173.11 | 81.157 | 2.365133039 | 1.04E-07 | 2.1136 |
| Hexacosanoic Acid | 446.02 | 59.3715 | 1.154365524 | 0.0071778 | 0.61389 |
| D-Glutamic Acid | 246.39 | 70.3786 | 2.160991554 | 1.22E-06 | 0.47834 |

## 4. Discussion

This pilot study provides the first multi-omics evidence suggesting that mothers of ARM-affected children harbor gut microbial and metabolomic signatures indicative of subclinical vascular dysfunction. The core findings include: (i) cases exhibited gut dysbiosis characterized by increased Firmicutes-to-Bacteroidetes ratio and reduced Coriobacteriaceae abundance; (ii) taxonomic analysis revealed significant enrichment of Faecalibacterium and depletion of Olsenella in cases; and (iii) metabolomic profiling identified four key alterations—D-glutamate depletion, thromboxane A2 elevation, reduced tungsten, and hexacosanoic acid reduction. These integrated findings suggest a preeclampsia-like maternal profile with distinct metabolomic and metagenomic signatures of vascular impairment. (Goulopoulou, 2017; Meijer et al., 2023; Youssef et al., 2022; Yusuf et al., 2001; Zhao et al., 2023). This study thus provides a potential mechanistic explanation for how maternal factors cause birth defects such as ARMs (Figure 9).

**Figure 9:**
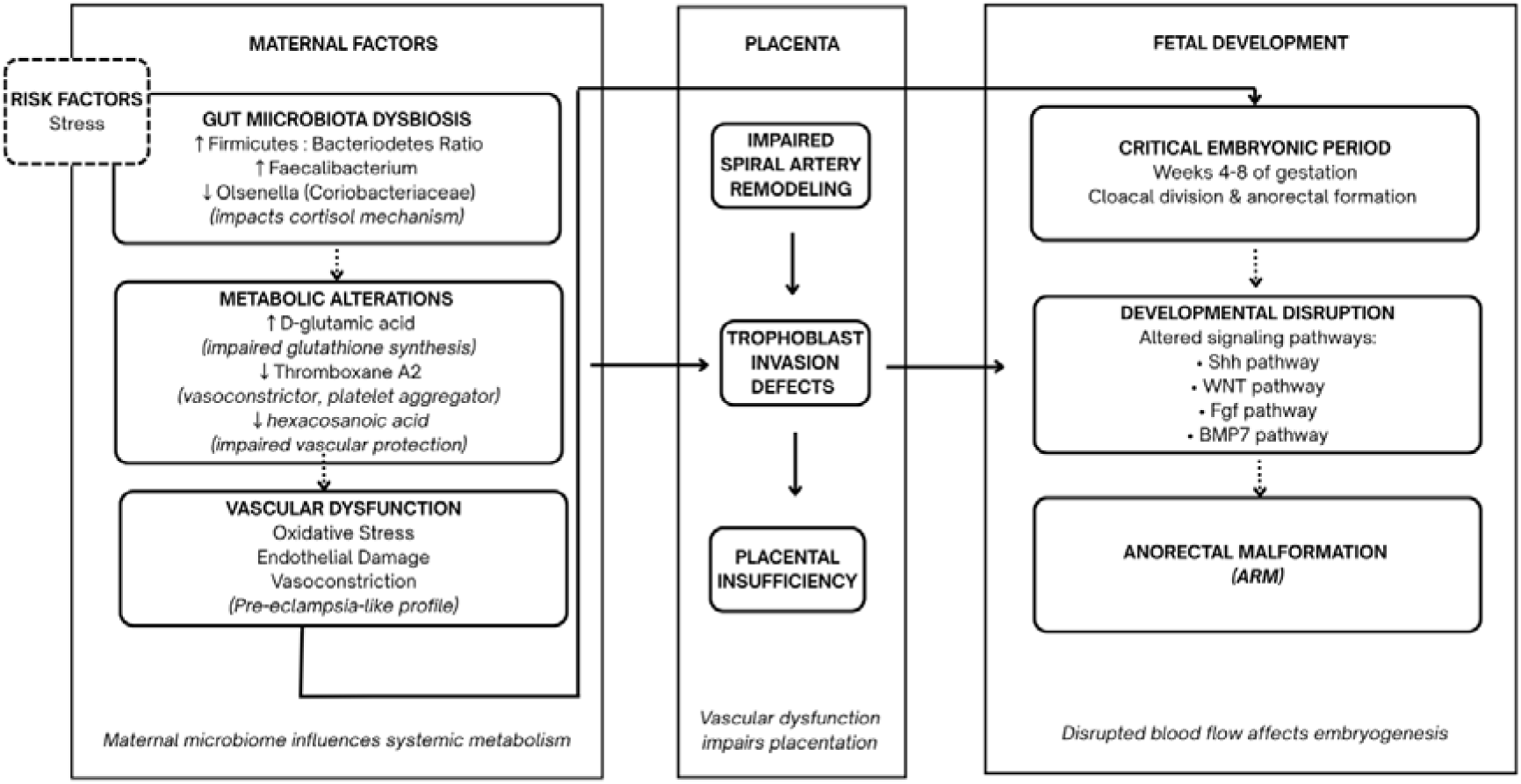
Proposed mechanism linking maternal vascular dysfunction to ARM development. Maternal stress and diet alter the gut microbiota, reflected in a higher Firmicutes to Bacteroidetes ratio and reduced Olsenella, a genus regulating cortisol metabolism. This dysbiosis, along with other physiological disruptions, leads to metabolic changes—lower D-glutamic acid, higher thromboxane A2, and reduced hexacosanoic acid—indicating impaired glutathione synthesis, increased vascular tone, and compromised peroxisomal function. These shifts signal oxidative stress and subclinical vascular injury, disrupting spiral artery remodeling and trophoblast

Metagenomic analysis revealed significant taxonomic differences in the gut microbiota of cases compared to controls. Although gut microbial diversity was slightly elevated, partial but distinct clustering in β-diversity suggested compositional differences. At the phylum level, cases exhibited a marked increase in Firmicutes and corresponding decrease in Actinobacteria. The elevated Firmicutes-to-Bacteroidetes ratio parallels patterns frequently observed in conditions associated with vascular dysfunction, including obesity, diabetes, and preeclampsia(Magne et al., 2020; Zhao et al., 2023).

A striking finding was the reduction in Coriobacteriaceae family (phylum Actinobacteria) in cases. These bacteria regulate steroid hormones—particularly host glucocorticoid metabolism. Within this family, Olsenella, a genus involved in bile acid transformation, showed depletion (S. Zhang et al., 2024). Since bile acids inhibit hepatic glucocorticoid clearance, the Olsenella deficit likely disrupts this regulatory axis, impairing cortisol catabolism and increasing circulating bioactive cortisol(McNeilly et al., 2010). Higher cortisol levels increase oxidative stress, impair uteroplacental blood flow, and are linked with hypertension and endothelial dysfunction—all involved in preeclampsia pathogenesis (Grzeszczak et al., 2023; Kosicka et al., 2018; F. Wang et al., 2014; D. Zhang et al., 2018).

Supporting this connection, Meijer et al. found depletion of Coriobacteriaceae in preeclampsia patients, while accruing evidence indicates that maternal stress or glucocorticoid exposure increases the risk of preeclampsia as well as various birth defects (Cottrell, 2009; Meijer et al., 2023; Talal AlSharif et al., 2023; Thalluri et al., 2022; Wu et al., 2022). Consistent with this pathway, we reported indicators of maternal psychological stress—a recognized inducer of cortisol production—in cases (Cottrell, 2009; Wu et al., 2022). Another notable observation at the genus level was significant enrichment of Faecalibacterium (phylum Firmicutes) in cases—a Gram-positive bacterium (Martín et al., 2023).

Metabolomic profiling identified four key alterations linked to vascular dysfunction in ARM pathogenesis. Disease enrichment analysis further supported this by revealing significant associations with coronary heart disease, adrenomyeloneuropathy, and neurovascular disorders—conditions sharing common vascular and oxidative stress pathways (Figure 9). First, we observed depletion of D-glutamic acid in cases. This D-glutamate enantiomer participates in a dynamic equilibrium with L-glutamate—a crucial substrate for glutathione production through the γ-glutamyl cycle (Chang et al., 2020; Di Girolamo et al., 2024). Reduced glutathione levels diminish protection against reactive oxygen species, leading to endothelial dysfunction and vascular impairment (Di Girolamo et al., 2024; Gutman & Parrilla, 2022). Accordingly, Youssef et al. reported aberrant D-glutamate and glutamate metabolism in early-onset severe Pre-eclampsia (Youssef et al., 2022).

Second, we discovered elevated thromboxane A2 (TXA2), a potent vasoconstrictor and platelet aggregator, indicating maternal vasoactive imbalance in cases(Goulopoulou, 2017). Beyond vasoconstriction, TXA2 disrupts placentation through multiple mechanisms: inhibiting trophoblast invasion and spiral artery remodeling, promoting trophoblast apoptosis, and impairing angiogenesis at the maternal-fetal interface(Gibbins et al., 2018;

Yusuf et al., 2001) (Figure 9). Third, we found reduced hexacosanoic acid (C26:0), a very long-chain saturated fatty acid (VLCFA) that undergoes β-oxidation primarily in peroxisomes and plays a role in endothelial protection(Lu et al., 2022; Weng et al., 2015). As VLCFAs play a crucial role in preventing pregnancy-induced hypertension, this depletion suggests compromised vascular resilience in cases(Li et al., 2020).

Fourth, cases showed reduced tungsten levels. Disease enrichment analysis (Table 4) linked this depletion to multiple sclerosis, Parkinson’s, and Alzheimer’s—conditions involving xanthine oxidase (XO)-mediated oxidative stress and cerebrovascular dysfunction (Biessels, 2022; Burrage et al., 2023; Geraldes et al., 2020; Haryuni et al., 2024; Higgins et al., 2009; Honorat et al., 2013; Kamaljeet et al., 2024). Notably, Tungsten demonstrates direct protective effects against vascular disease through xanthine oxidase inhibition. For example, Tungsten treatment prevents tumor necrosis factor-α (TNF-α)-induced injury of brain endothelial cells through inhibition of xanthine oxidase activity (Terada et al., 1992). This is particularly relevant to preeclampsia, where elevated TNF-α drives endothelial dysfunction via similar oxidative stress pathways (Roberts & Hubel, 2009). Tungsten also prevents atherosclerosis development in ApoE knockout mice by reducing vascular superoxide formation, preventing endothelial dysfunction, and attenuating inflammatory gene expression (Schröder et al., 2006). Thus, tungsten depletion may compromise endothelial protection against XO-mediated injury, increasing oxidative vascular damage.

Collectively, these findings indicate that even in the absence of overt vascular disease, mothers of ARM-affected children exhibit subclinical vascular dysfunction. This dysfunction, likely driven by alterations in gut microbiota and serum metabolites, may compromise placental development and perfusion during the critical window of cloacal division in early gestation. We propose a mechanistic model wherein maternal vascular health—modulated by microbial composition and metabolic imbalance—disrupts key developmental signaling pathways (e.g., Sonic hedgehog, Wnt, FGF), thereby increasing the risk of anorectal malformations (Figure 9). The discovery of specific microbial and metabolic profiles linked to ARMs presents opportunities for early risk identification and prevention. By targeting maternal vascular health therapeutically, both preconceptionally and in early pregnancy, it may be possible to reduce ARM incidence and improve outcomes for affected families.

### Limitations and Future Work

Several limitations of this study exist. The cross-sectional case-control design cannot establish causality or temporal sequence between maternal vascular dysfunction and ARM development. Although our results suggest plausible biological mechanisms, longitudinal studies beginning pre-conceptionally or in early pregnancy are necessary to confirm whether these maternal signatures precede ARM formation. Postpartum sample collection may not fully reflect maternal physiology during the critical first-trimester window of anorectal development. While gut microbial communities and certain metabolic parameters show relative stability, first-trimester sampling would provide more direct evidence of conditions present during organogenesis. Despite identifying significant differences in key parameters, our sample size limits statistical power for subgroup analyses and exploration of potential confounders. Additionally, direct measurement of placental function and maternal vascular parameters would strengthen our proposed mechanistic pathway. Finally, whether elevated cortisol directly drives the observed metabolic alterations requires further investigation as potential links between cortisol dysregulation, thromboxane A2 production, and peroxisomal function could reveal a unified pathophysiological cascade rather than isolated findings. Future research should address these limitations through prospective designs, larger cohorts, and direct vascular function assessment to validate our findings.

## Conclusion

This study offers new insight into the pathogenesis of anorectal malformations by implicating maternal vascular dysfunction as a potentially modifiable contributor. Through integrated microbiome and metabolomic analysis, we identify a molecular profile—characterised by Olsenella depletion, altered glutamate metabolism, elevated thromboxane A2, and reduced peroxisomal lipid markers—that signals subclinical impairment in vascular tone, oxidative balance, and endothelial function. In this light, ARMs may arise not solely from genetic aberrations, but from transient physiological conditions in early pregnancy. While preliminary, these findings opens the possibility that maternal health—specifically vascular stability—may be monitored, supported, and even intervened upon before structural anomalies take form. Thus, our findings lay the groundwork for a broader research agenda that shifts the focus from postnatal repair to antenatal prevention.

## Abbreviations

ALD: Adrenoleukodystrophy
ARMs: Anorectal malformations
ASVs: Amplicon sequence variants
BMP7: Bone morphogenetic protein 7
CHDs: Congenital heart defects
DEMs: Differentially expressed metabolites
DNA: Deoxyribonucleic acid
EC: Enzyme Commission
ESI+: Electrospray ionization (positive mode)
FC: Fold change
FDR: False discovery rate
FGF: Fibroblast growth factor
HMDB: Human Metabolome Database
IQR: Interquartile range
KEGG: Kyoto Encyclopedia of Genes and Genomes
LC-MS: Liquid chromatography-mass spectrometry
LC/Q-TOF: Liquid chromatography/quadrupole time-of-flight
OPLS-DA: Orthogonal partial least squares discriminant analysis
OTU: Operational taxonomic unit
PCA: Principal component analysis
PCoA: Principal coordinate analysis
PCR: Polymerase chain reaction
PERMANOVA: Permutational multivariate analysis of variance
QIIME2: Quantitative Insights Into Microbial Ecology version 2
rRNA: Ribosomal ribonucleic acid
Shh: Sonic hedgehog
TNF-α: Tumor necrosis factor-alpha
TXA2: Thromboxane A2
VIP: Variable importance in projection
VLCFA: Very long-chain fatty acids
Wnt: Wingless-related integration site
XO: Xanthine oxidase

## Author Contributions

The study was conceptualized and designed by **B.P.**, who also led funding acquisition, ethical approvals, data interpretation, graphics, metagenomics data analysis, and contributed to both the original draft and manuscript revisions. **A.M.** provided overall supervision and led funding acquisition, sample collection and processing, data curation, metabolomics data analysis, and methodology writing, while also contributing to manuscript review and editing. **R.J.** and **J.R.** led study design, ethical approvals, supervision, and clinical data collection, and contributed to the review and editing of the manuscript. **A.K.** led data analysis and draft compiling, and provided supporting contributions in sample collection, isolation, and preparation. **S.V.** led sample collection and processing and supported sample analysis. **S.S.** supported sample collection, processing, and analysis. **K.S.** led sample collection and processing and contributed to sample analysis. **V.B.** provided support in departmental resource coordination and contributed in manuscript reviewing. All authors read and approved the final manuscript.

## Data Availability Statement

The raw 16S rRNA gene sequencing data will be deposited in the NCBI Sequence Read Archive (SRA), and metabolomics data (including raw files, processed peak tables, and associated metadata) will be submitted to MetaboLights. Accession numbers will be provided upon publication.

## Ethical approval

Applicable (EC/Approval/104/2023/04/12/2023) from the Institutional Ethics Committee (IEC) of B.J. Medical College & Civil Hospital, Ahmedabad

## Informed consent

Applicable

## Data availability statement

Not applicable

## Acknowledgement

None

---

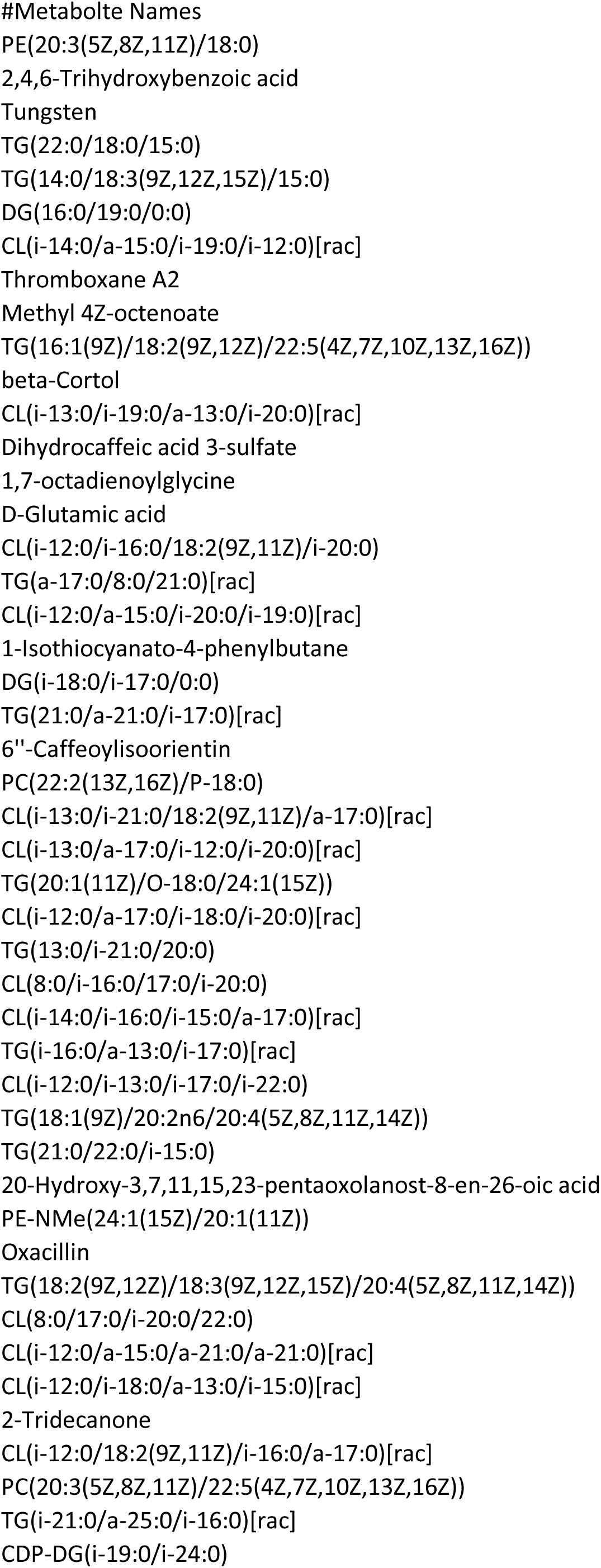
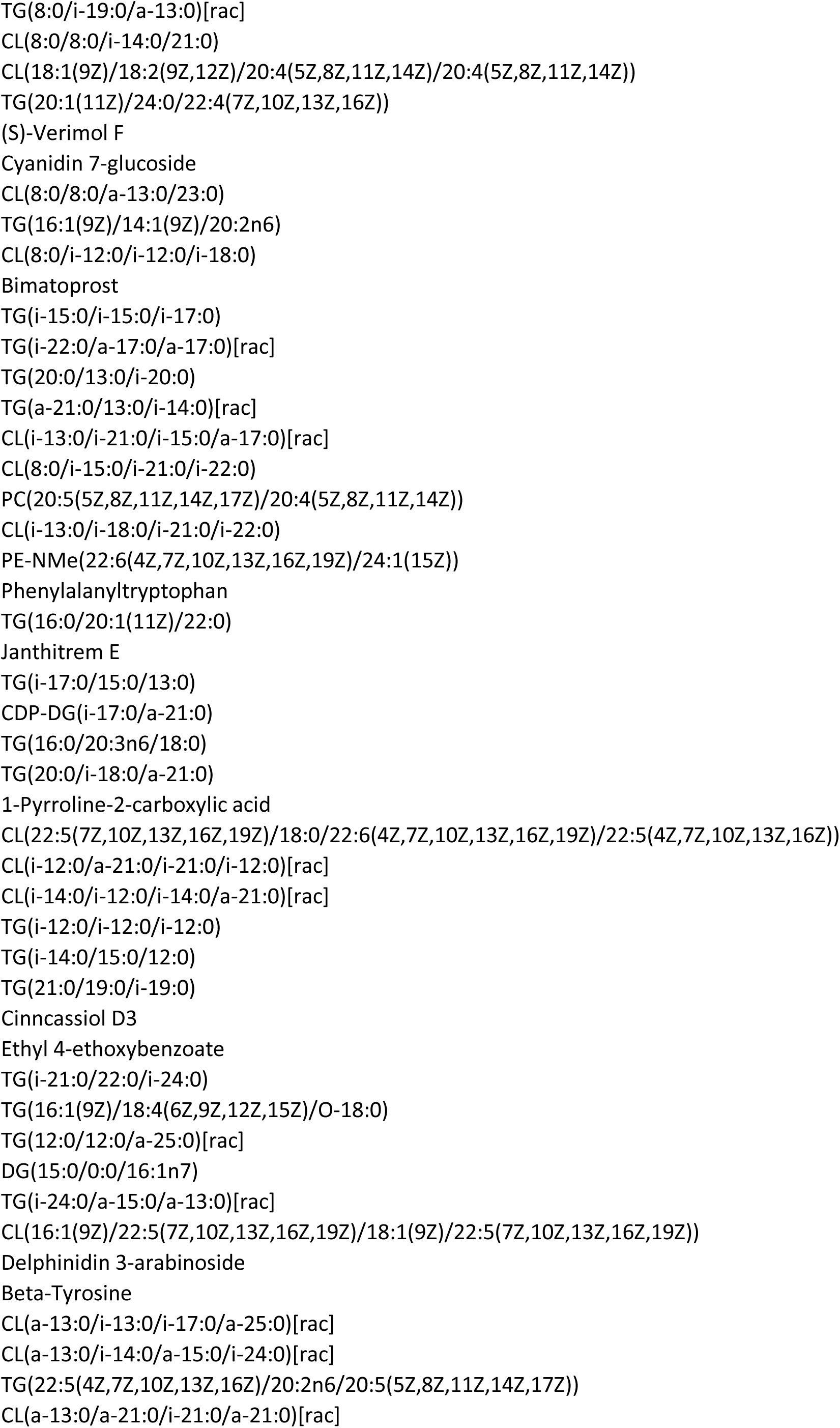
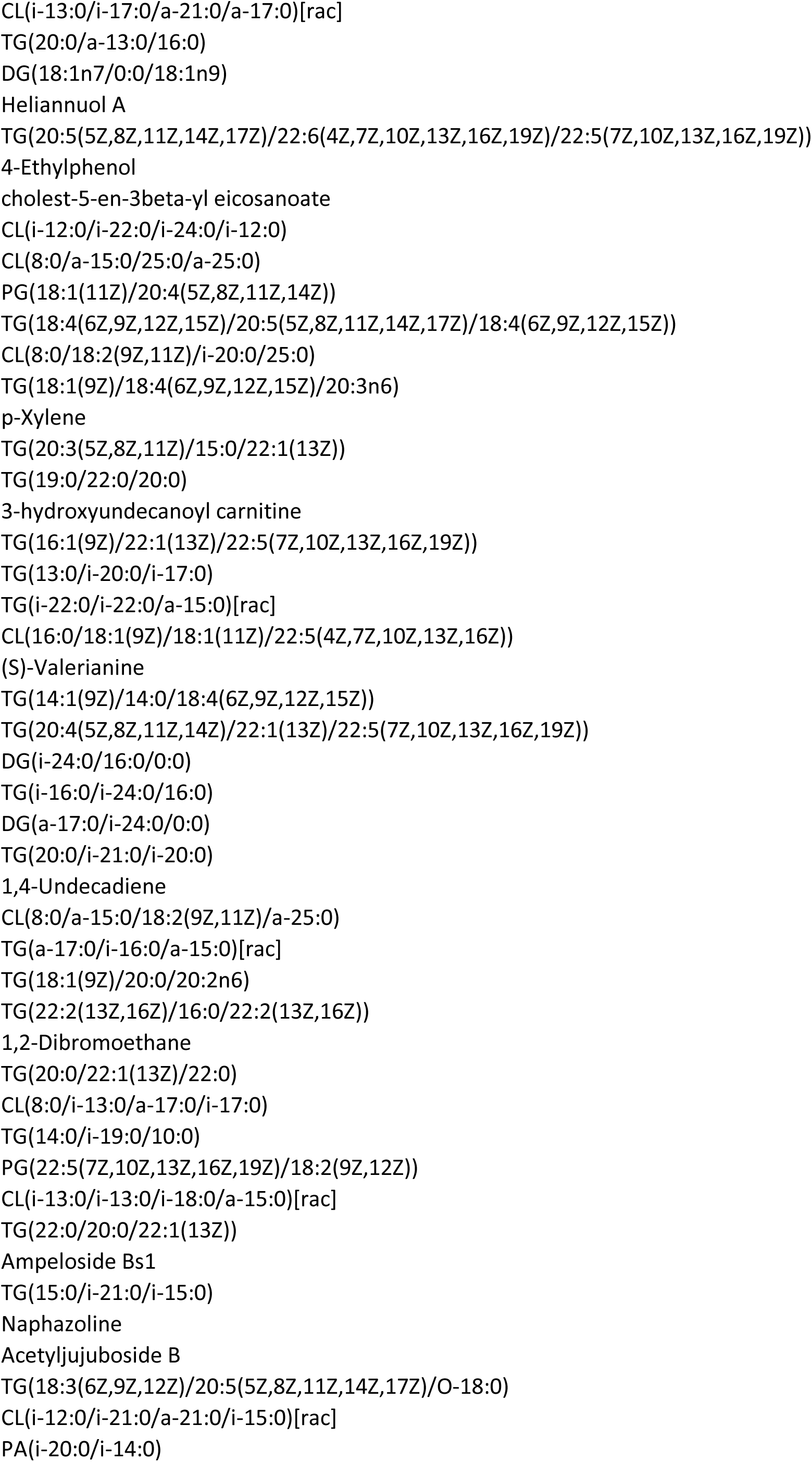
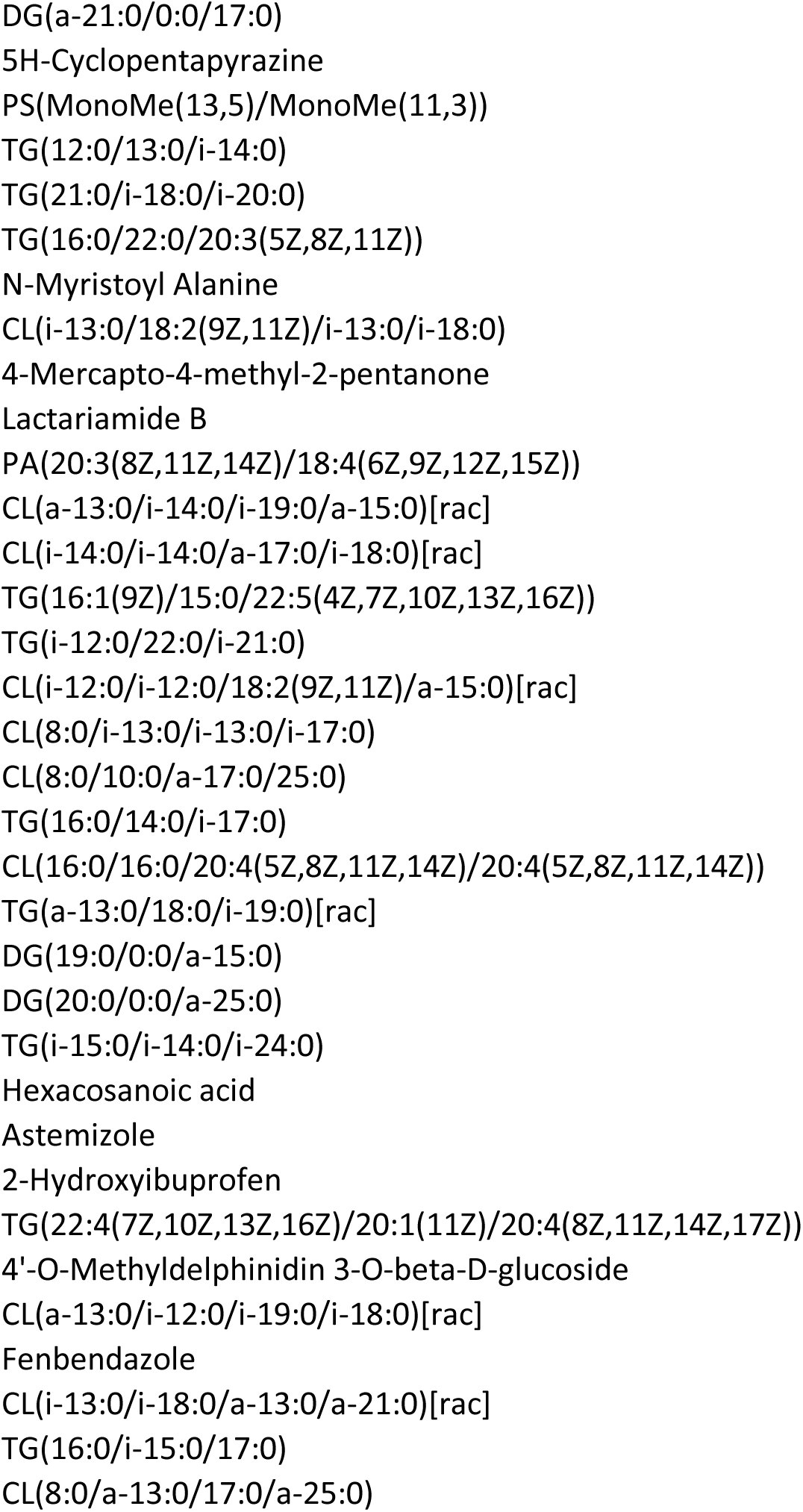

**Supplementary Table 1:** Demultiplexed sequence read statistics. This table summarizes the minimum, median, mean, maximum, and total number of reads obtained for both forward and reverse sequencing reads across all samples.

|  | forward reads | reverse reads |
| --- | --- | --- |
| Minimum | 151446 | 151446 |
| Median | 359802 | 359802 |
| Mean | 492627 | 492627 |
| Maximum | 1022477 | 1022477 |
| Total | 9852532 | 9852532 |

**Supplementary Figure 1 :**
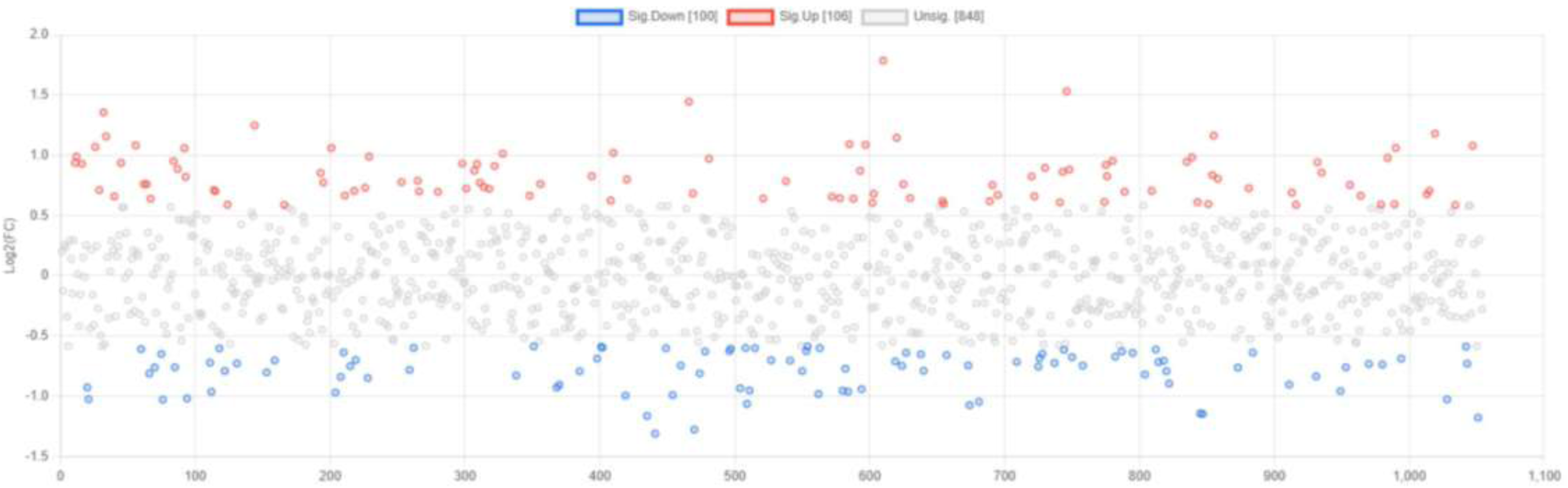
Fold change plot for individual features, indicating significantly upregulated and downregulated mass features between case and control groups. Points colored in blue represent significantly down-regulated genes (n=100), red points indicate significantly up-regulated genes (n=106), and grey points denote genes that are not significantly differentially expressed (n=848).

